# Meta-Analysis and Systematic Review of the Measures of Psychological Safety

**DOI:** 10.1101/2024.02.09.24302562

**Authors:** Jenny JW Liu, Natalie Ein, Rachel A. Plouffe, Julia Gervasio, Kate St. Cyr, Anthony Nazarov, J. Don Richardson

**Author notes:** Address for correspondence: Jenny JW Liu, PhD, MacDonald Franklin Operational Stress Institute Research Centre.

## Abstract

**Purpose:** In a psychologically safe environment, individuals feel safe to share thoughts, acknowledge errors, experiment with new ideas, and exhibit mutual respect. However, there is little consensus on how psychological safety should be measured and the constructs that make up psychological safety. This meta-analysis and systematic review sought to evaluate the quality of measures used to assess psychological safety.

**Methodology:** The meta-analysis and systematic review were conducted using Cochrane’s guidelines as a framework for data synthesis. A total of 217 studies were included in this review.

**Findings:** Across 217 studies, the average internal consistency value ranged from Cronbach’s alpha of .77 to .81, with considerable heterogeneities across samples (I2 = 99.92, Q[221] = 259632.32, p < .001). Together, findings suggest that the quality of existing measures evaluating psychological safety may be acceptable.

**Originality:** There is room for improvement with respect to examinations of factor structures within psychological safety, the degree of association between psychological safety and other constructs, and opportunities for exploring similarities and differences across populations and contexts.

## Introduction

Psychological safety has been referred to as one’s perceived security within an organization such that they feel safe to take interpersonal risks, voice opinions, and share ideas without fear of recourse [1–3]. There has been an increase in both academic and industry interest in psychological safety due to the associated benefits at both micro (individual and interpersonal) and macro (organizational) levels. Research has demonstrated how support from leaders and organizations foster psychologically safe environments [4–7]. When an individual feels psychologically safe, they are more likely to share thoughts, acknowledge errors, experiment with new ideas, and exhibit mutual respect for other team members, and less likely to experience burnout [1, 8]. Subsequently, psychologically safe environments facilitate greater information sharing, learning behaviours (e.g., seeking ways to improve processes), and team performance [2, 6, 9]. Ultimately, organizations with high psychological safety exhibit decreased employee turnover [10], increased innovation [11], and increased overall performance [12].

Researchers have continued to develop self-report measures to assess individuals’ perceptions of psychological safety in their workplace. For example, a significant portion of the extant literature measures psychological safety with the original or an adapted version of the Edmondson’s Psychological Safety Scale (PSS) [1]. However, it is important to consider that the conceptualization of psychological safety and the context under which the scale was developed may have evolved since its introduction in 1999. The PSS measures an individual’s perception of team level psychological safety as a unidimensional construct and at the time of scale development, researchers’ understanding of psychological safety was largely focused on individuals’ perceived ability to take risks in the workplace [2]. In retrospect, this narrow conceptualization may limit the potential to evaluate a wider range of constructs which may contribute to feelings of psychological safety. Other constructs may include supportive leaders [6], clear expectations of individual and organizational roles and values [9], and cohesive climate, environment, or network [2]. As this literature has evolved, multiple varying definitions and associated measures of psychological safety have emerged resulting in a lack of clear consensus on how psychological safety should be measured and the constructs that make up psychological safety. It is unclear whether these constructs serve as factors that mutually reinforce psychological safety, represent distinct components within a larger umbrella of psychological safety, or are simply correlated yet distinct constructs from psychological safety.

This lack of consensus in our understanding of psychological safety also perpetuates challenges for practical application. For example, it is unclear whether our current understandings of psychological safety is adequately captured with existing measures. Further, the reliability and validity of these measures remains largely understudied. To date, no research has systematically identified and compiled all existing psychological safety measures and subsequently assessed the quality of these measurement tools. With this in mind, we conducted a meta-analysis and systematic review of the literature to evaluate the reliability and validity of all existing measures reflecting psychological safety across various population characteristics (e.g., military, civilian). Specifically, we sought to: (1) identify all existing psychological safety measures in the literature, (2) examine their overall reliability and validity, and (3) assess the quality of these measures in capturing psychological safety.

## Methods

The meta-analysis and systematic review were conducted using Cochrane’s guidelines as a framework for data synthesis [13]. This review was not registered, and a protocol was not prepared. SWIFT-Active Screener, a web-based, collaborative review software, was used to accelerate the screening processes [14]. Specifically, SWIFT enlists a proprietary machine learning algorithm to prioritize relevant articles for screening with a high degree of accuracy [14]. Using this approach, our review and screening times were reduced with no risks or losses to accuracy of screening.

### Search Terms

A search of the literature was conducted on October 26, 2021. Five databases were used: PsycINFO (OVID), MEDLINE (OVID), PsycTests-ProQuest, SCOPUS, and ProQuest Dissertations & Theses. The following search terms were used: safety, psychological safety, team, learning, ethics, training, processes, effectiveness, dynamics, organization, identification, culture, leadership, inclusion, climate, feedback environment, climate for inclusion, ethics training, military, veterans, service members, soldiers, first responders, military ethics, military teams (see Supplementary Material for string terms).

### Inclusion and Exclusion Criteria

The inclusion criteria consisted of studies with adult human populations, studies that examined psychological safety, and studies implementing a measurement tool capturing psychological safety. The exclusion criteria consisted of review articles, non-empirical work (e.g., book chapters), and studies that measured psychological safety, but did not include any data analysis (e.g., theory-driven articles).

### Study Selection and Data Extraction

At each review stage, citations were reviewed by two independent raters. Ten raters were engaged to complete this study. Interrater reliability via the percentage accuracy of screeners’ selections were high at each stage (title and abstract at 82.7% and full text review at 82.5%).

Conflict resolution was conducted as a group until complete consensus was acquired. A total of 217 studies with 238 independent samples for psychological safety measures and 368 independent samples for the convergent measures were included in this review (see Figure 1).

**Fig 1.**
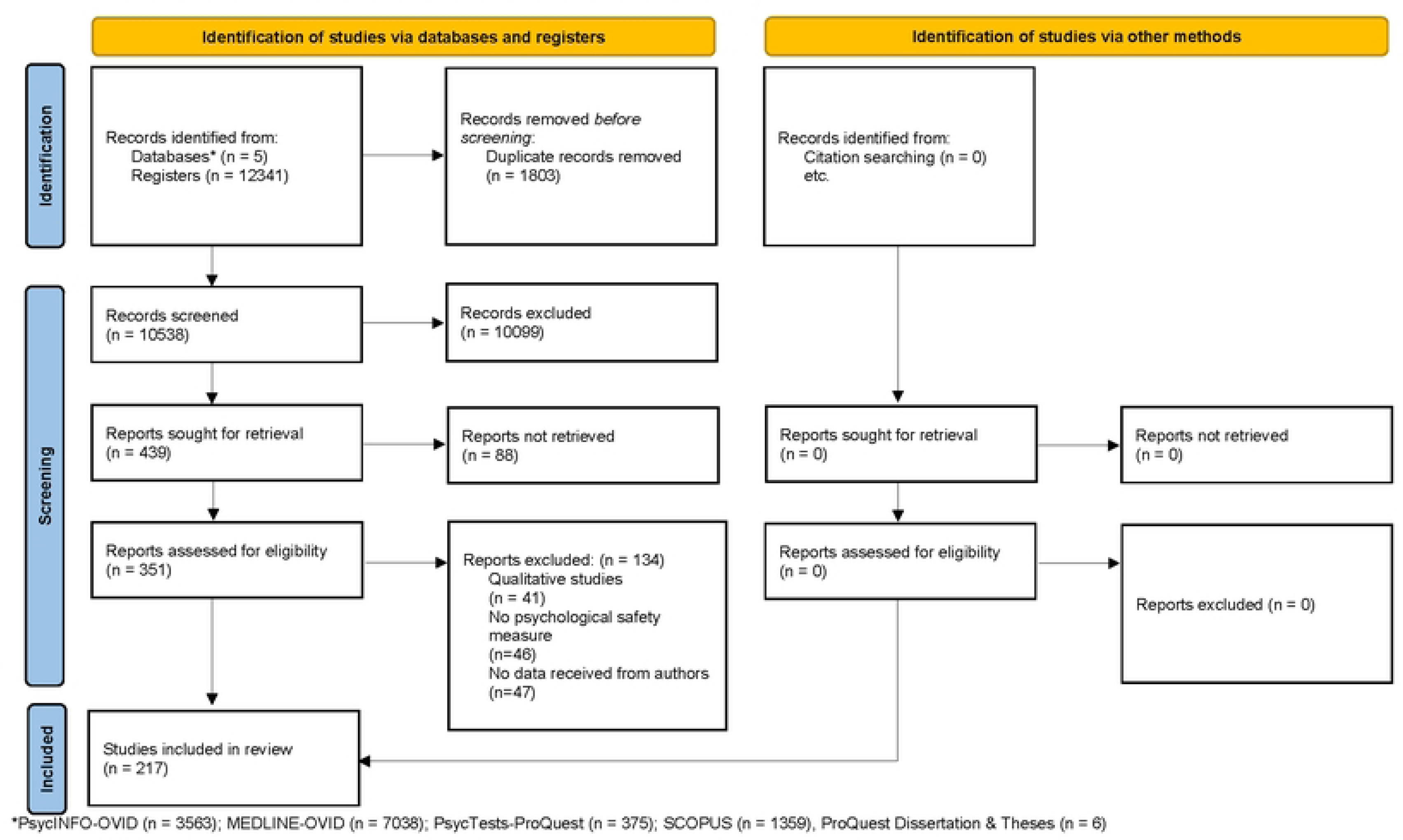
Preferred Reporting Items for Systematic Reviews (PRISMA) Flow Diagram [15]

The following information was extracted from each article: study characteristics, psychological safety measurement tool features, psychometric properties of the measurement tool, and convergent measure features. The study characteristics included: (1) *sample population* used in the study (military, paramilitary, civilian, or mixed), (2) *sample size*, and (3) *percentage of men* in the study sample.

The psychological safety measurement tool features included: (1) *name of the psychological safety measure* used Edmondson’s [PSS] [1], Liang *et al*.’s Psychological Antecedents of Promotive and Prohibitive Voice [PAPPV] [16], May *et al*.’s Measure of Psychological Conditions of Meaningfulness, Safety, and Availability, and the Engagement of the Human Spirit at Work [MPCMSAE] [5], Carmeli *et al*.’s Psychological Safety Measure [PSM] [17], Detert and Burris’s Psychological Safety Measure [PSM] [18], or other [a variety of psychological safety scales that were used less than 5 times across studies], (2) *number of scale anchors* (if applicable; e.g., 5-point Likert type), (3) *number of items* in each measure, (4) *version of the measure* (original or modified), (5) *translation of the measure* (not translated or translated), and (6) whether the research *involved a measurement validation study* (yes or no).

The psychometric properties of the measurement tool included: (1) *means and standard deviations of the measure*, (2) *internal consistency* (Cronbach’s alpha), (3) *test-retest reliability* (if applicable), and (4) whether the researchers *performed a factor analysis* (yes or no) or (5) regression (yes or no).

The convergent measure features included: (1) *number of measures used*, (2) *construct measured* (validated psychological safety measure, team climate, performance, leadership, risk taking, trust, organizational belonging, job satisfaction or support), (3) *measure name*, (4) *author of the measure*, (5) *version of the measure* (original or modified), and (6) *correlation magnitude and direction* with the psychological safety measure.

### Analytic Strategy for Meta-Analysis

Data analyses were conducted using Statistical Package for the Social Sciences (SPSS) Software Version 27 [19] and Comprehensive Meta-Analysis (CMA) software Version 3 [20]. Primary analyses included evaluations of the internal consistency of psychological safety measures via Cronbach’s alphas relative to population groups, measurement tool features, and psychometric properties. The sampling distribution across studies was computed using the means, standard deviations, and sample sizes. The relative influence of study population characteristics and measurement tool features were then examined against the computed sampling distribution. The convergent validity of psychological safety measures in relation to related constructs were examined using correlations. Finally, publication bias was assessed via visual inspection of the funnel plot, Egger’s Regression Test, Durval and Tweedie’s trim-and-fill, and classic fail-safe to determine the strength and confidence of evidence in the literature.

### Study Rigour for Systematic Review

To assess the quality of the psychological safety measures used in these studies, we assessed study rigour for each sample. The studies were organized based on the psychological safety scale used (i.e., Edmondson’s PSS [1], Liang *et al*.’s PAPPV [16], May *et al*.’s MPCMSAE [5], Carmeli *et al*.’s PSM [17], Detert and Burris’s PSM [18], or other). We adopted the Consensus-Based Standards for the Selection of Health Measurement Instruments (COSMIN) definition and criteria of evaluation to determine study rigour [21]. For each study, the following information was used to determine study rigour score: 1) scale reliability (i.e., the values of Cronbach alpha and/or test-retest reliability), 2) whether the measure used was an original scale or a modified version, 3) factorial/criterion validity (i.e., whether any hypothesis testing using these statistical approaches were implemented: a scale validation study, performed a factor analysis and/or performed a regression predicting a criterion variable, 4) whether the convergent measures were original and/or modified, and 5) scale translations (i.e., if applicable, how the psychological safety scale was translated into another language).

To determine study rigour, we assigned each category a numerical value (see Table 1). After scoring each category, the scores were summed and an overall rigour score was assigned. The following overall rigour scores were applied to studies without scale translation data: 5 or less (poor), 6 to 8 (fair), 9 to 11 (good) and 12 and above (excellent). If the study involved translation of a scale, one point was added to each cut-off threshold (i.e., overall scoring was categorized as follows: 6 or less [poor], 7 to 9 [fair], 10 to 12 [good] and 13 and above [excellent]).

**Table 1.** Study Rigour Scoring Criteria.

| Categories | Numerical Value |  |  |  |
| --- | --- | --- | --- | --- |
|  | 1 | 2 | 3 | 4 |
| **Reliability** | $\alpha \leq .69$ | $\alpha = .70$ to $.79$ | $\alpha = .80$ to $.89$ | $\alpha \geq .90$ *or*<br>Test-Retest Reliability |
| **Original vs Modified** | Modified *or*<br>Combined Scales | Original | -- | -- |
| **Factorial/Criterion**<br>**Validity** | No Hypothesis Testing | 1 Statistical Approach | $\geq 2$ Statistical Approaches<br>*or* Validation Study Only | Validation Study *and* $\geq 1$<br>Statistical Approach |
| **Convergent Measures** | None | Modified | Original *and* Modified | Original |
| **Scale Translation**<br>**(when applicable)** | Not Ideal | Ideal | -- | -- |

**Notes.** Not ideal = did not report how the scale was translated; Ideal = back translation or outlining how many raters were used to translate the scale.

## Results

Across the 217 articles, the sample contained data from 504,136 individuals (average of 53% men), including 224 studies with civilians (94.1%), 8 studies with military members (3.4%), two studies with paramilitary populations in the form of Public Safety Personnel (0.8%), and four studies with mixed populations (1.7%; see Table S1 for raw data of included articles). Psychological safety was evaluated using self-report measures in all studies. A large majority (*n* = 130, 54.6%) employed Edmondson’s PSS to measure psychology safety [1]. Of the remaining studies, 9.7% (*n* = 23) used Liang *et al*.’s PAPPV [16], 7.6% (*n* = 18) used May *et al*.’s MPCMSAE [5], 3.4% (*n* = 8) used Carmeli *et al*.’s PSM [17], 2.1% (*n* = 5) used Detert and Burris’s PSM [18], and 22.7% (*n* = 54) used other (i.e., alternative self-report measures or a combination of scale items from multiple measures). The number of scale items varied from two to 38. Items were scored using Likert-type scales ranging from 4 points to 10 points. The most frequently used Likert-type scoring was a 5-point Likert scale (61.8%, *n* = 139; see Table 2).

**Table 2.**
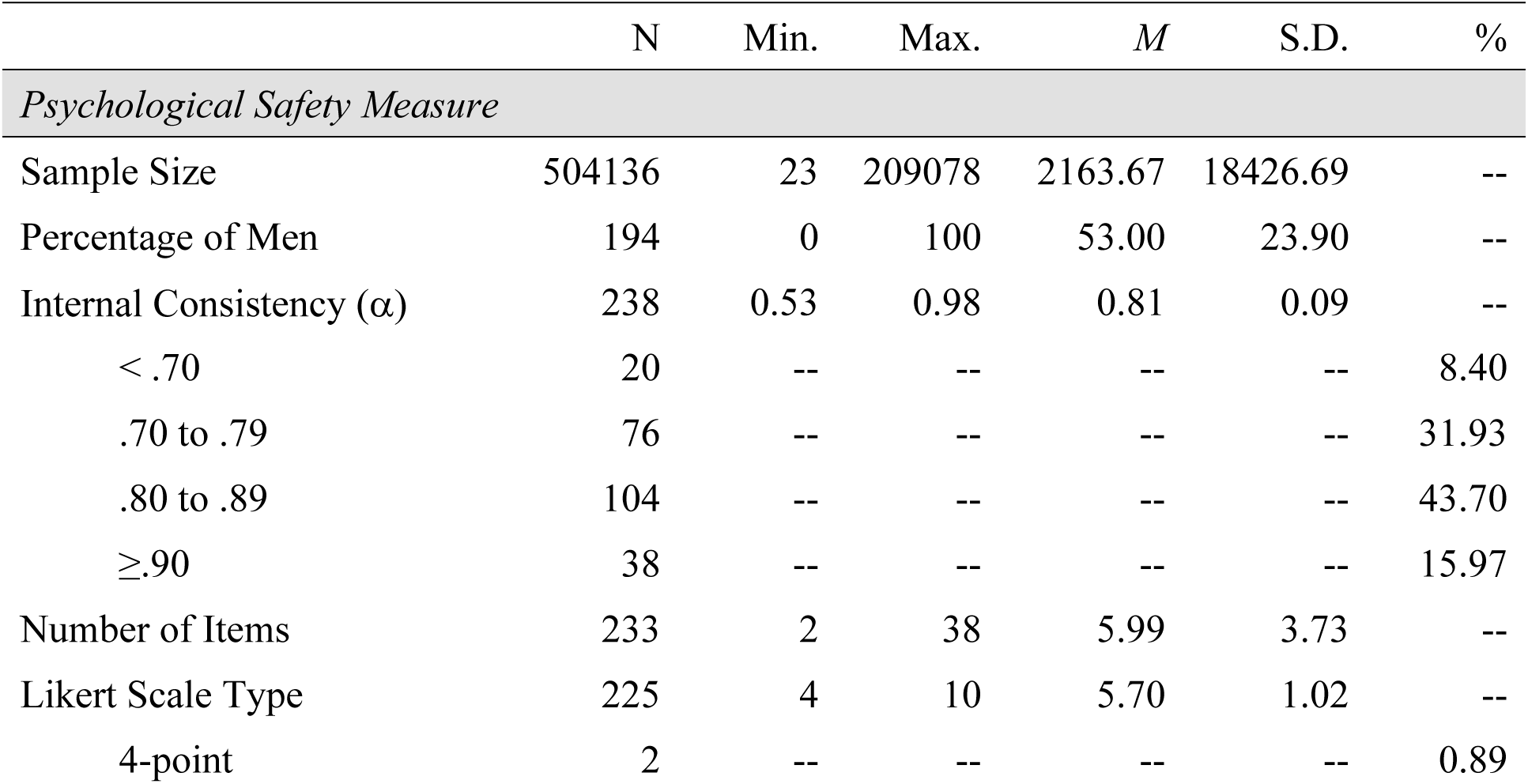
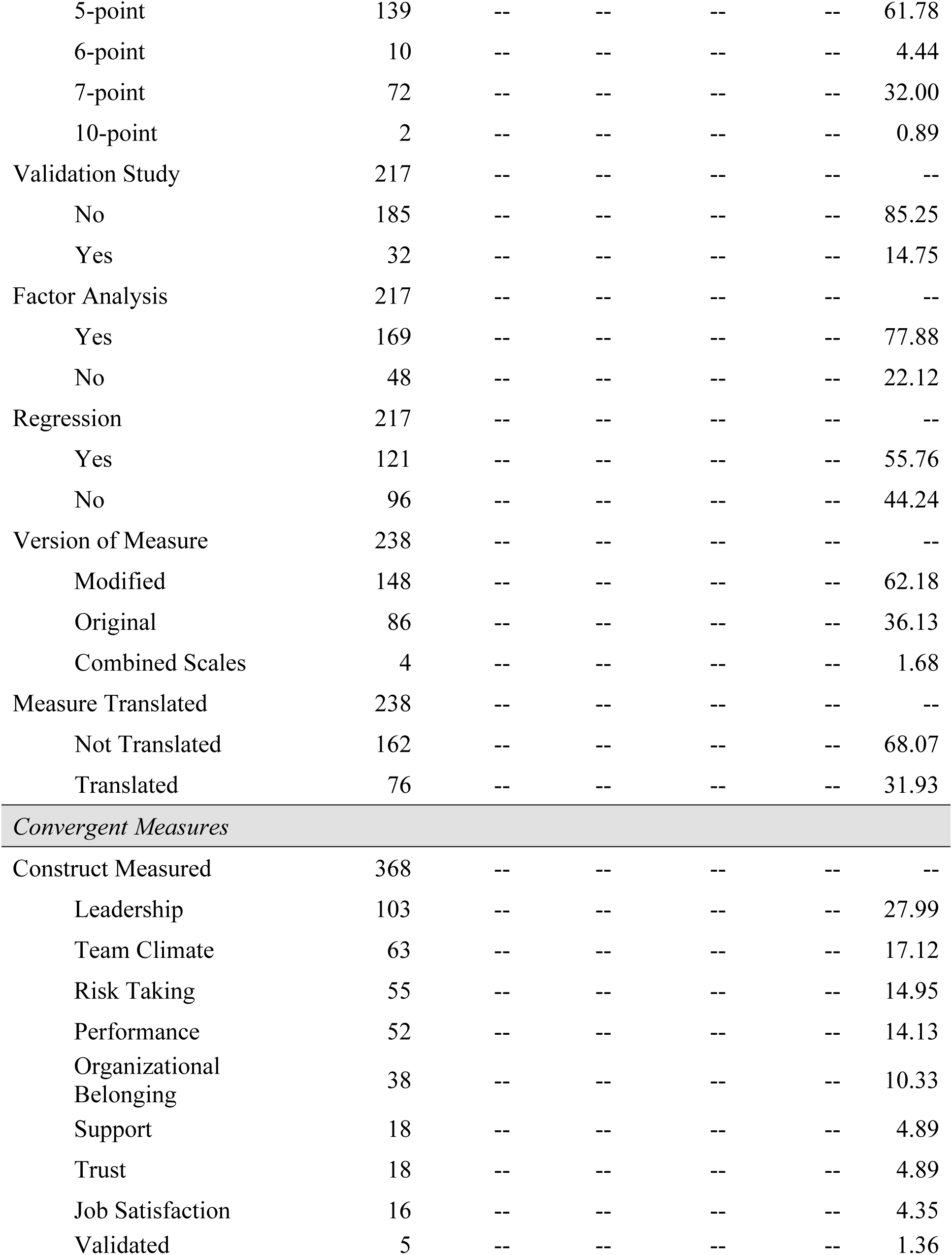
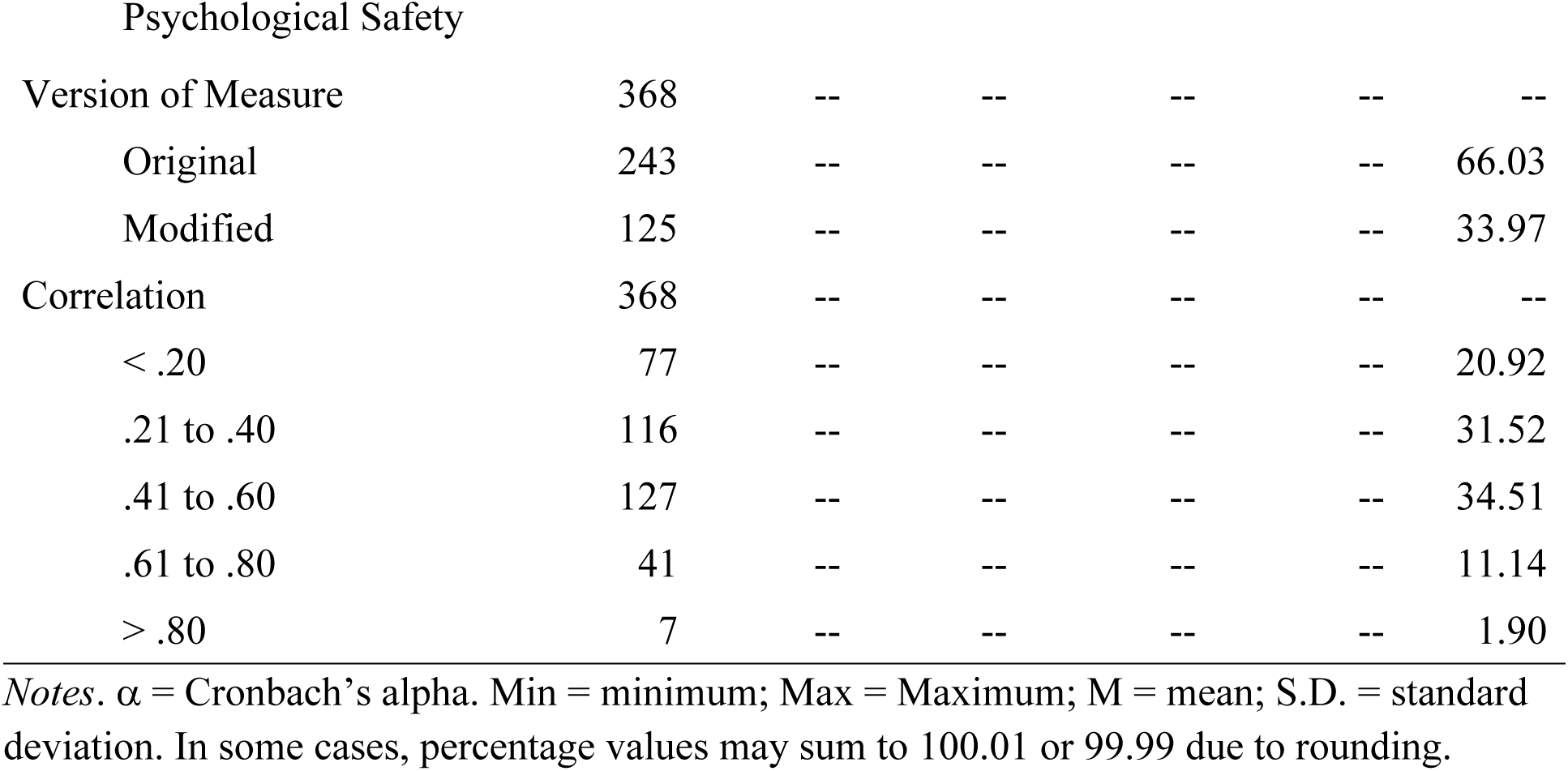
Characteristics of Included Studies.

|  | N | Min. | Max. | *M* | S.D. | % |
| --- | --- | --- | --- | --- | --- | --- |
| *Psychological Safety Measure* |  |  |  |  |  |  |
| Sample Size | 504136 | 23 | 209078 | 2163.67 | 18426.69 | -- |
| Percentage of Men | 194 | 0 | 100 | 53.00 | 23.90 | -- |
| Internal Consistency ($\alpha$) | 238 | 0.53 | 0.98 | 0.81 | 0.09 | -- |
| < .70 | 20 | -- | -- | -- | -- | 8.40 |
| .70 to .79 | 76 | -- | -- | -- | -- | 31.93 |
| .80 to .89 | 104 | -- | -- | -- | -- | 43.70 |
| $\geq .90$ | 38 | -- | -- | -- | -- | 15.97 |
| Number of Items | 233 | 2 | 38 | 5.99 | 3.73 | -- |
| Likert Scale Type | 225 | 4 | 10 | 5.70 | 1.02 | -- |
| 4-point | 2 | -- | -- | -- | -- | 0.89 |
| 5-point | 139 | -- | -- | -- | -- | 61.78 |
| 6-point | 10 | -- | -- | -- | -- | 4.44 |
| 7-point | 72 | -- | -- | -- | -- | 32.00 |
| 10-point | 2 | -- | -- | -- | -- | 0.89 |
| Validation Study | 217 | -- | -- | -- | -- | -- |
| No | 185 | -- | -- | -- | -- | 85.25 |
| Yes | 32 | -- | -- | -- | -- | 14.75 |
| Factor Analysis | 217 | -- | -- | -- | -- | -- |
| Yes | 169 | -- | -- | -- | -- | 77.88 |
| No | 48 | -- | -- | -- | -- | 22.12 |
| Regression | 217 | -- | -- | -- | -- | -- |
| Yes | 121 | -- | -- | -- | -- | 55.76 |
| No | 96 | -- | -- | -- | -- | 44.24 |
| Version of Measure | 238 | -- | -- | -- | -- | -- |
| Modified | 148 | -- | -- | -- | -- | 62.18 |
| Original | 86 | -- | -- | -- | -- | 36.13 |
| Combined Scales | 4 | -- | -- | -- | -- | 1.68 |
| Measure Translated | 238 | -- | -- | -- | -- | -- |
| Not Translated | 162 | -- | -- | -- | -- | 68.07 |
| Translated | 76 | -- | -- | -- | -- | 31.93 |
| *Convergent Measures* |  |  |  |  |  |  |
| Construct Measured | 368 | -- | -- | -- | -- | -- |
| Leadership | 103 | -- | -- | -- | -- | 27.99 |
| Team Climate | 63 | -- | -- | -- | -- | 17.12 |
| Risk Taking | 55 | -- | -- | -- | -- | 14.95 |
| Performance | 52 | -- | -- | -- | -- | 14.13 |
| Organizational<br>Belonging | 38 | -- | -- | -- | -- | 10.33 |
| Support | 18 | -- | -- | -- | -- | 4.89 |
| Trust | 18 | -- | -- | -- | -- | 4.89 |
| Job Satisfaction | 16 | -- | -- | -- | -- | 4.35 |
| Validated | 5 | -- | -- | -- | -- | 1.36 |

REVIEW OF THE MEASURES OF PSYCHOLOGICAL SAFETY
| Psychological Safety |  |  |  |  |  |  |
| --- | --- | --- | --- | --- | --- | --- |
| Version of Measure | 368 | -- | -- | -- | -- | -- |
| Original | 243 | -- | -- | -- | -- | 66.03 |
| Modified | 125 | -- | -- | -- | -- | 33.97 |
| Correlation | 368 | -- | -- | -- | -- | -- |
| < .20 | 77 | -- | -- | -- | -- | 20.92 |
| .21 to .40 | 116 | -- | -- | -- | -- | 31.52 |
| .41 to .60 | 127 | -- | -- | -- | -- | 34.51 |
| .61 to .80 | 41 | -- | -- | -- | -- | 11.14 |
| > .80 | 7 | -- | -- | -- | -- | 1.90 |

*Notes.* $\alpha$ = Cronbach's alpha. Min = minimum; Max = Maximum; M = mean; S.D. = standard deviation. In some cases, percentage values may sum to 100.01 or 99.99 due to rounding.

### Internal Consistency and Sampling Distribution

The psychometric properties of psychological safety across samples were examined primarily through evaluations of internal consistency (Cronbach’s alphas) and sampling distributions. Across 238 samples, the average Cronbach’s alpha was reported as *a* = .81, which represents good internal consistency of constructs of psychological safety. In other words, the psychological safety measures generally assessed a homogeneous construct, as anticipated. Average Cronbach’s alphas were reported as follows across measures: *a* = .77 (May *et al*.’s MPCMSAE) [5], *a* = .80 (Edmondson’s PSS) [1], *a* = .83 (Liang *et al*.’s PAPPV) [16], *a* = .84 (Carmeli *et al.*’s PSM) [17], and *a* = .84 (Detert and Burris’s PSM) [18].

Further examinations of the range of alphas indicated that over 75% of studies reported a Cronbach’s alpha of between .70 and <.90, while 16.0% reported Cronbach’s alphas in the high range of .90+. Univariate analyses of variance examined differences in internal consistency across scales, and there were significant differences across measures (*F*[5, 232] = 2.61, *p* = .026). Specifically, studies using Liang *et al*.’s PAPPV [16] measure reported significantly higher internal consistencies relative to studies using May *et al.*’s MPCMSAE [5], *p* = .021 (see Figure 2).

**Fig 2.**
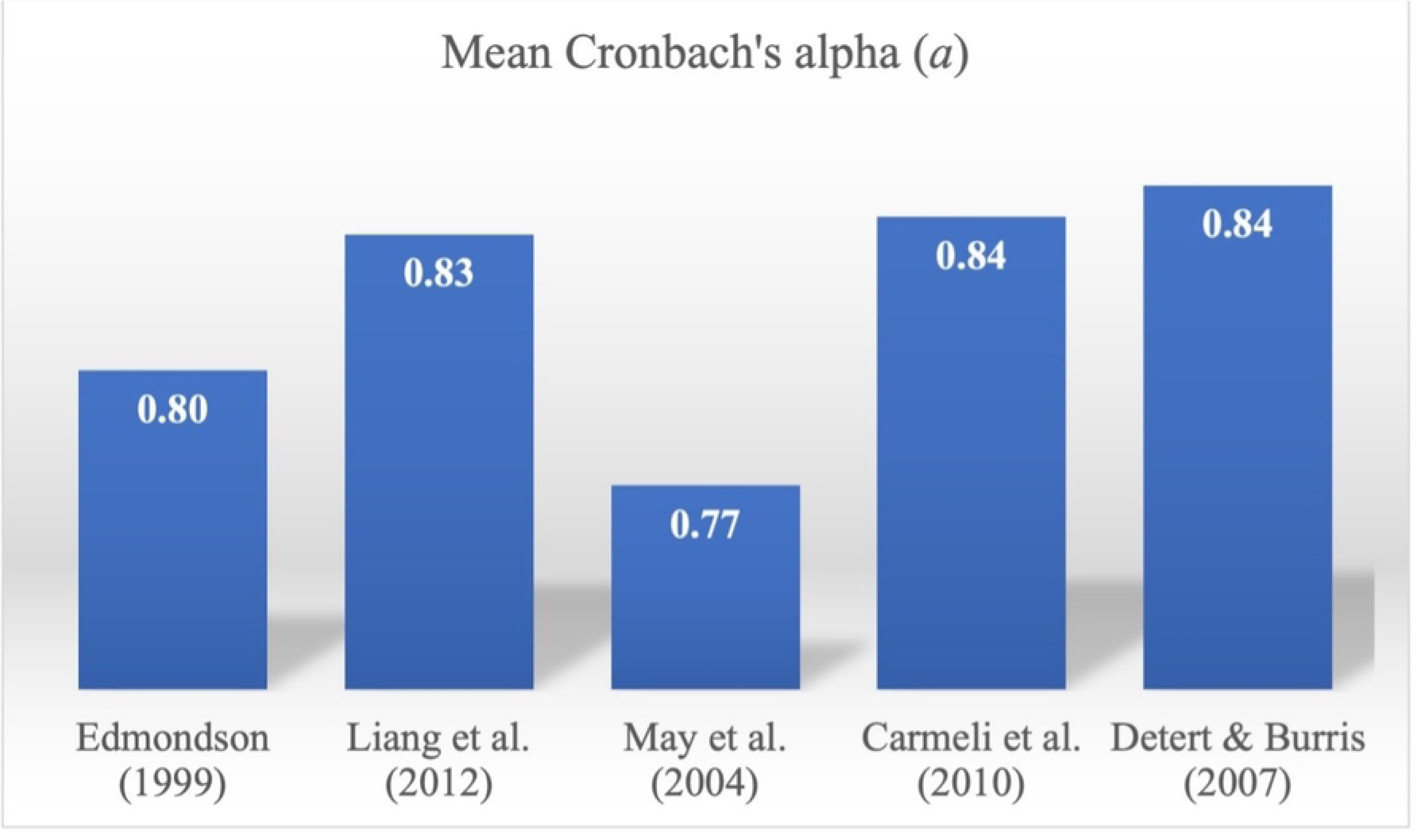
Mean Cronbach’s alpha of Psychological Safety Measures

The point estimates were significantly different across measures of psychological safety, *Q*(5) = 93.09, *p* < .001. May *et al*.’s MPCMSAE [5] reported the highest (point estimate = 6.14, *SE* = 0.43, 95% confidence interval [CI] = 5.29, 6.98), followed by Edmondson’s PSS [1] (point estimate = 4.67, *SE* = 0.10, 95% confidence interval [CI] = 4.48, 4.86), other (point estimate = 4.33, *SE* = 0.06, 95% confidence interval [CI] = 4.20, 4.45), Detert and Burris’s PSM [18] (point estimate = 4.07, *SE* = 0.22, 95% confidence interval [CI] = 3.64, 4.50), Liang e*t al.*’s PAPPV [16] (point estimate = 3.81, *SE* = 0.09, 95% confidence interval [CI] = 4.48, 4.86), and Carmeli *et al.*’s PSM [17] (point estimate = 3.64, *SE* = 0.12, 95% confidence interval [CI] = 3.42, 3.86). Given the lack of military samples for each of the measures, point estimates were not compared between civilians and non-civilians for each of the measures. Instead, an overall comparison found that effects were significantly different across samples of civilians (point estimate = 4.63, *SE* = 0.07, 95% confidence interval [CI] = 4.50, 4.76), and military populations (point estimate = 7.39, *SE* = 0.70, 95% CI [6.02, 8.77]), *Q*(3) = 116.92, *p* < .001.

A meta-regression using the moment methods estimator for random effects was used to examine the degree of influence of Cronbach’s alpha, Likert-type anchors, and the number of items in the measures may have on the overall sampling distributions of psychological safety measures. Together, these factors accounted for a significant proportion of between-study variance (*r^2^* = .13). Indeed, all factors emerged as significant predictors in the model. While Cronbach’s alpha significantly negatively predicted sampling distribution (*b* = −2.12, *Z* = −3.59, *p* = .003), Likert-type (*b* = 0.88, *Z* = 16.42, *p* < .003) and number of items in the measure (*b* = 0.09, *Z* = 4.96, *p* < .003) both positively predicted sampling distribution.

### Test-Retest Reliability

Of the studies included, only six reported test-retest reliability, which represents the stability of the tested construct over time. Of these studies, test-retest reliability coefficients ranged from .27 (poor) to .98 (excellent). An average taken across samples indicated the overall test-retest reliability was .66, suggesting good stability in the overall construct of psychological safety [22]. Additional forms of psychometric properties were infrequently conducted across studies. Of the included studies, 85.3% (*n* = 185) did not involve forms of scale validation, 22.1% (*n* = 48) did not examine the factor structure of the scales used, and 44.2% (*n* = 96) did not conduct a regression analysis to evaluate criterion validity.

### Convergent Measures

The convergent validity of various psychological safety measures was examined using available correlations with related constructs. Across measures, correlations with psychological safety ranged from *r* = -.58 to *r* = .90, with an absolute mean correlation of *r* = .38. A univariate analysis of variance examined differences in the absolute values of correlations across construct domains. Tests of between-subjects effects found no significant differences in correlations across construct domains *F*(8, 335) = 1.49, *p* = .16, *n^2^* = .03. Due to low sample sizes across psychological safety measures and related constructs, analyses were not conducted between measures (see Table 3 for correlations between psychological safety measures and each construct domain; see Table S2 for raw data of convergent measures).

**Table 3.** Mean Correlations between Psychological Safety Measures and Convergent Measures and their Related Constructs.

|  | **Edmondson's PSS**<br>**(1999)** | **Liang *et al.*'s**<br>**PAPPV (2012)** | **May *et al.*'s**<br>**MPCMSAE (2004)** | **Carmeli *et al.*'s**<br>**PSM (2010)** | **Detert and Burris's**<br>**PSM (2007)** |
| --- | --- | --- | --- | --- | --- |
|  | *Mean Corr. (k)* | *Mean Corr. (k)* | *Mean Corr. (k)* | *Mean Corr. (k)* | *Mean Corr. (k)* |
| Job Satisfaction | .30 (7) | -- | .42 (2) | -- | -- |
| Leadership | .29 (55) | .39 (12) | .26 (8) | .50 (6) | .63 (3) |
| Organizational Belong | .40 (14) | .48 (5) | .30 (7) | .61 (1) | .14 (3) |
| Performance | .35 (34) | .28 (2) | .21 (6) | .51 (3) | -- |
| Risk-Taking | .24 (22) | .39 (18) | .39 (5) | -- | .30 (4) |
| Support | .47 (5) | .51 (2) | -- | .27 (1) | -- |
| Team Climate | .36 (39) | .41 (3) | .18 (4) | -- | .32 (1) |
| Trust | .29 (8) | .51 (5) | .29 (1) | -- | .16 (1) |
| Validated PS | .31 (3) | -- | -- | -- | -- |

*Note.* Mean Corr. = mean correlation; PSS = Psychological Safety Scale; PAPPV = Psychological Antecedents of Promotive and Prohibitive Voice; MPCMSAE = Measure of Psychological Conditions of Meaningfulness, Safety, and Availability, and the Engagement of the Human Spirit at Work; PSM = Psychological Safety Measure; PS = Psychological Safety. -- = no convergent measures associated with this psychological safety measure. The most common psychological safety scales and related convergent measures are reported. Given that the "other" category encompasses multiple different psychological safety measures, correlations could not be meaningfully interpreted and therefore are not reported here.

### Publication Bias

Visual inspection of the funnel plot suggests that there may be some imputed missing studies to the left of the center, but they are unlikely to influence the overall effects observed in our sample (see Figure 3). Egger’s Test of the Intercept reported an intercept (B0) of 12.70, 95% confidence interval (7.86, 17.54), with *t* = 5.17, *df* = 231. The 1-tailed *p*-value (recommended) is 0.00, and the 2-tailed *p*-value is 0.00. Using Duval and Tweedie’s Trim and Fill procedure, analyses suggested that 64 studies are missing. Under the random effects model, the point estimate for the combined studies was 4.50 (95% confidence interval [CI] = 4.48, 4.66). Using Trim and Fill, the imputed point estimate was 3.64 (95% CI = 3.54, 3.74). This review incorporated data from 233 studies, which yielded a *z*-value of 1808.13 and corresponding 2-tailed *p*-value of 0.00. The fail-safe *N* is 198 297 972. This means that there would need to be 851064.3 missing studies for every observed study for the effect to be nullified.

**Fig 3.**
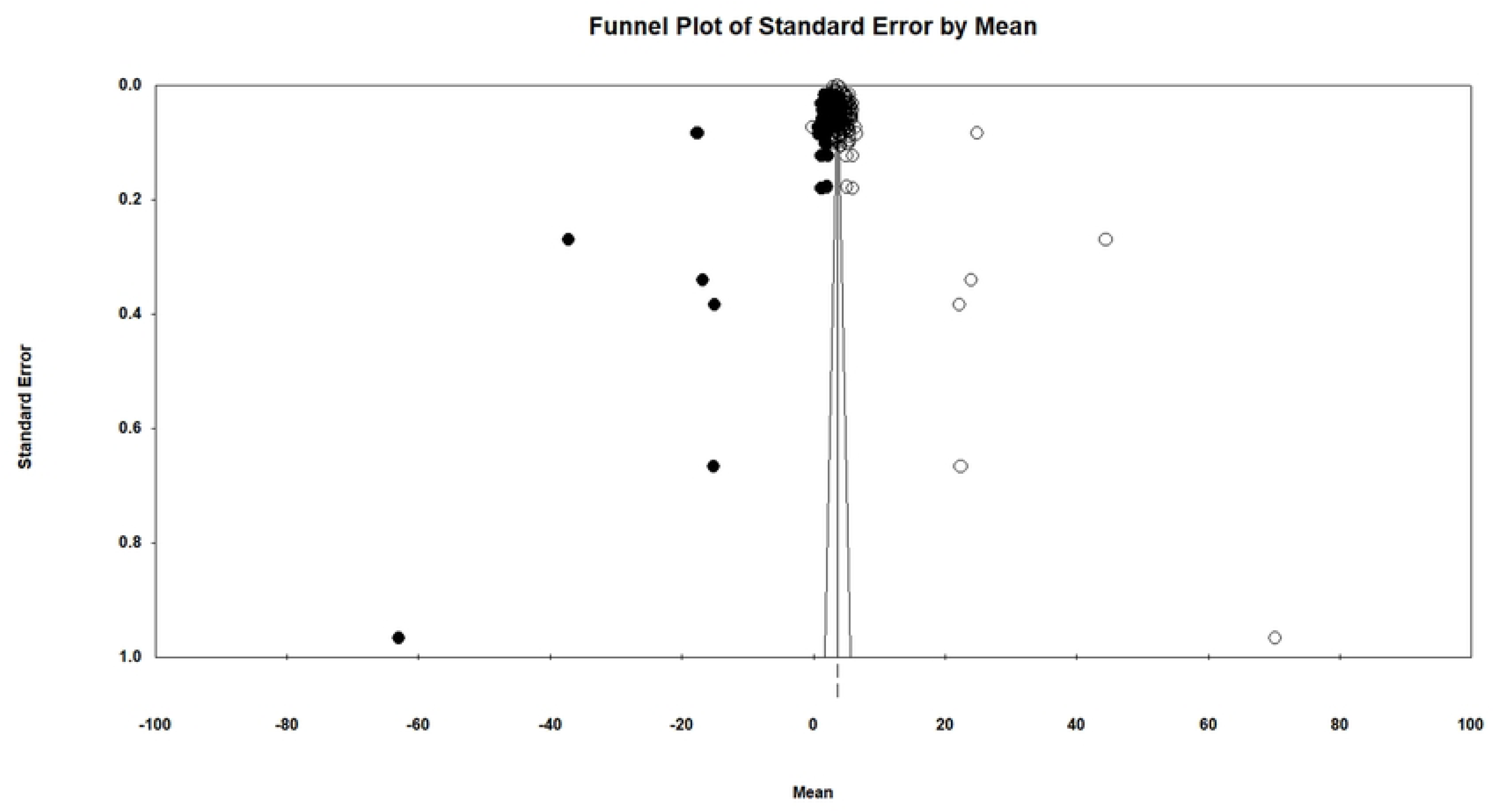
Funnel Plot

### Study Rigour Results

Overall, the majority of scales were rated as “good” (see Table 4). Across all scales and categories of evaluation for study rigour, cross-cultural translations (where applicable) emerged as the most robust category, with the highest average score across studies, followed by use of convergent measures, and use of original versus modified measures. Interestingly, reliability assessments via Cronbach’s alpha and factorial/criterion validity testing emerged as the categories with the lowest average rigour rating (see Table S3 and S4 for raw data).

**Table 4.**
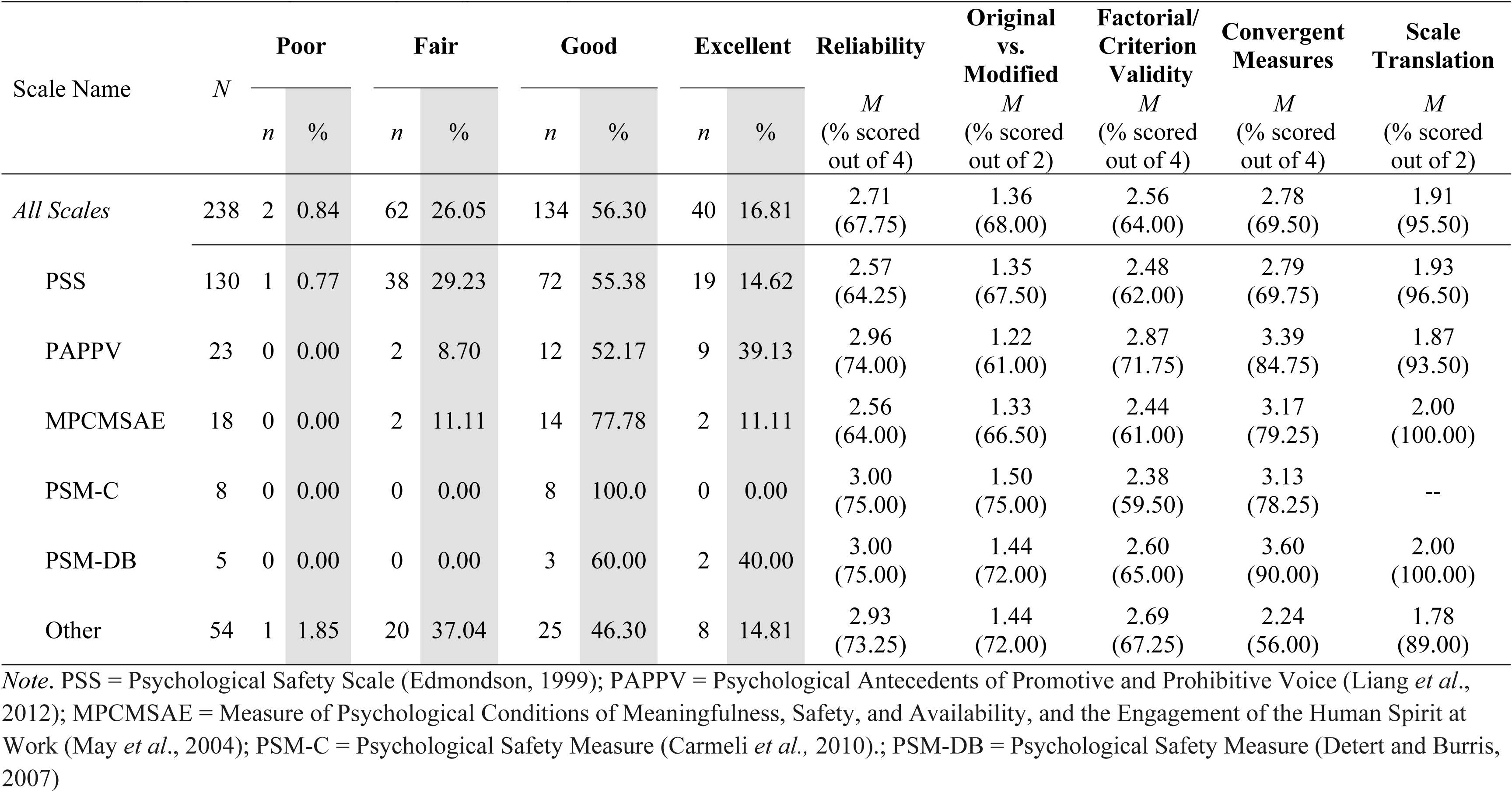
Study Rigour Rating across Psychological Safety Measures.

| Scale Name | *N* | Poor |  | Fair |  | Good |  | Excellent |  | Reliability | Original vs. Modified | Factorial/ Criterion Validity | Convergent Measures | Scale Translation |
| --- | --- | --- | --- | --- | --- | --- | --- | --- | --- | --- | --- | --- | --- | --- |
|  |  | *n* | % | *n* | % | *n* | % | *n* | % | *M*<br>(% scored out of 4) | *M*<br>(% scored out of 2) | *M*<br>(% scored out of 4) | *M*<br>(% scored out of 4) | *M*<br>(% scored out of 2) |
| *All Scales* | 238 | 2 | 0.84 | 62 | 26.05 | 134 | 56.30 | 40 | 16.81 | 2.71<br>(67.75) | 1.36<br>(68.00) | 2.56<br>(64.00) | 2.78<br>(69.50) | 1.91<br>(95.50) |
| PSS | 130 | 1 | 0.77 | 38 | 29.23 | 72 | 55.38 | 19 | 14.62 | 2.57<br>(64.25) | 1.35<br>(67.50) | 2.48<br>(62.00) | 2.79<br>(69.75) | 1.93<br>(96.50) |
| PAPPV | 23 | 0 | 0.00 | 2 | 8.70 | 12 | 52.17 | 9 | 39.13 | 2.96<br>(74.00) | 1.22<br>(61.00) | 2.87<br>(71.75) | 3.39<br>(84.75) | 1.87<br>(93.50) |
| MPCMSAE | 18 | 0 | 0.00 | 2 | 11.11 | 14 | 77.78 | 2 | 11.11 | 2.56<br>(64.00) | 1.33<br>(66.50) | 2.44<br>(61.00) | 3.17<br>(79.25) | 2.00<br>(100.00) |
| PSM-C | 8 | 0 | 0.00 | 0 | 0.00 | 8 | 100.0 | 0 | 0.00 | 3.00<br>(75.00) | 1.50<br>(75.00) | 2.38<br>(59.50) | 3.13<br>(78.25) | -- |
| PSM-DB | 5 | 0 | 0.00 | 0 | 0.00 | 3 | 60.00 | 2 | 40.00 | 3.00<br>(75.00) | 1.44<br>(72.00) | 2.60<br>(65.00) | 3.60<br>(90.00) | 2.00<br>(100.00) |
| Other | 54 | 1 | 1.85 | 20 | 37.04 | 25 | 46.30 | 8 | 14.81 | 2.93<br>(73.25) | 1.44<br>(72.00) | 2.69<br>(67.25) | 2.24<br>(56.00) | 1.78<br>(89.00) |

*Note.* PSS = Psychological Safety Scale (Edmondson, 1999); PAPPV = Psychological Antecedents of Promotive and Prohibitive Voice (Liang *et al.*, 2012); MPCMSAE = Measure of Psychological Conditions of Meaningfulness, Safety, and Availability, and the Engagement of the Human Spirit at Work (May *et al.*, 2004); PSM-C = Psychological Safety Measure (Carmeli *et al.*, 2010).; PSM-DB = Psychological Safety Measure (Detert and Burris, 2007)

When evaluating the categories of rigour across the use of specific psychological safety measures, several deviations from the overall trends can be observed. Detert and Burris’s PSM [18] was observed to have higher than average rigour ratings across all categories despite less frequent use across studies. Conversely, while Edmondson’s PSS [1] was the most frequently used measure of psychological safety, it had lower than average rigour ratings in three out of five categories while the remaining two categories were only on par with average ratings. The remaining three scales (Liang *et al*.’s PAPPV [16], May *et al*.’s MPCMSAE [5], and Carmeli *et al.*’s PSM [17]) exhibited more variability in scoring across categories (e.g., May *et al.*’s MPCMSAE [5] scored lower than average on three categories, but higher than average in two categories). Finally, all psychological safety scales examined here implemented translations in some studies, except for Carmeli *et al.*’s PSM [17], whereby no published studies adopted the measure for translation.

## Discussion

This meta-analysis and systematic review sought to identify and assess the quality of existing psychological safety measures. Overall, the quality of psychological safety measures were good, with some areas identified as needing further improvement.

Across all psychological safety measures, the Cronbach’s alphas fell within the “good” range, with scale items consistently capturing a homogenous construct. Less than 20% of the measures evaluated scored in the high range which may initially indicate room for improvement; however, it is also plausible that Cronbach’s alpha values over .90 may reflect redundant items. Importantly, outside of evaluations of internal consistency, there was little evidence to support the reliability and validity of measures of psychological safety. Specifically, evaluations of test-retest reliability, factorial, and criterion validity were identified as major gaps in the literature. Our evaluations of study rigour found that Detert and Burris’s PSM [18] was the most rigorous scale despite the low representative sample, while Edmondson’s PSS [1] was found to be the least rigorous scale. Importantly, samples using Edmondson’s PSS [1] made up more than half (55%) of all included samples and was likely overrepresented relative to other common scales and drove the overall trend in ratings.

Despite its widespread use, Edmondson’s measure may not be as contemporarily useful as it was constructed to reflect team psychological safety as a unidimensional construct. Indeed, Edmondson and Lei [2] indicated that there should be a shift in our understanding of psychological safety from a unidimensional team-level construct to a multidimensional interpersonal construct. Specifically, psychological safety reflects multiple factors, such as climate of trust and respect. In evaluations of convergent validity measures, the diverse domains (e.g., job satisfaction, team climate) typically yielded a low correlation (averaging *r* = .39). Our findings based on convergent measures suggest that existing measures of psychological safety may not be strongly related to domains that contribute to a more modern conceptualization of psychological safety. We also identified areas of consideration which may inform the development of standardized measurement tools. For example, over half of the studies reviewed used a 5-point Likert-type scale for scoring. In addition, we found reliability and factorial and criterion validity to have the lowest scores in study rigour, despite being among the most important dimensions of psychometric evaluations. Thus, future efforts at scale development should focus on confirming factor structures, in addition to examining multiple indexes of reliability and validity.

Across all studies, psychological safety was mostly measured in civilian populations, with less than 5% of overall studies examining psychological safety in military and paramilitary settings. Psychological safety is considered a critical factor in performance and teamwork [2]. This can be especially relevant in military settings, where the ability to perform successfully can mitigate negative outcomes (e.g., injury, death). Indeed, research has identified psychological safety to be a strong predictor of team performance in military units [23]. Specifically, feeling safe, trusting each other, communicating effectively, and cooperation, were all vital to the success of various missions and objectives.

Taken together, meta-analytic and systematic review evidence on the quality of studies suggests that while existing measures are acceptable, more work is needed to advance the study of psychological safety in line with evolving understandings of the construct and scientific rigour. Scale development efforts and evaluations of psychological safety across contexts and populations should consider the potential multidimensional nature of the construct.

### Strength of Evidence and Future Considerations

In addition to examinations of the quality of measurement tools, we also tested for the presence of publication bias, or the degree to which published findings may be influenced by the results found in this sample. Indicators of publication bias suggest the presence of some publication biases that may have skewed the results to be more positive. However, these biases are minor. Analyses of publication biases thus suggest the findings from the current study can be interpreted with confidence.

Although the overall reliability and validity of measures was adequately assessed, specific characteristics of the measures, such as the number of scale items and the use of 5-point Likert-type ratings should be adopted and applied to the design and development of new psychological safety scales. Finally, our findings suggest the presence of differences in psychological safety based on the distribution of data across populations of civilians and military/paramilitary samples. These findings suggest that more research is needed to examine psychological safety through a multidimensional lens, with more rigorous assessments of reliability and validity, and in a wider variety of populations and contexts.

## Data Availability

All relevant data are within the manuscript and its Supporting Information files.

## Acknowledgement

We would like to thank the following individuals who have contributed to this report: Bailey Wolfs, Chayanika Tyagi, Clara Baker, Dac Hung Nguyen, Janissha Pushpaharan, Jasper Crockford, Jessica Zhang, Juwon Omar, Karmen Tse, Samdarsh Saroya, Suriya Ragu, and Valerie Kulakouski.

## Competing Interests

The author(s) declare none.

## Notes

### Competing Interest Statement

The authors have declared no competing interest.

### Funding Statement

The author(s) received no specific funding for this work.

